# Data-driven Testing Program Improves Detection of COVID-19 Cases and Reduces Community Transmission

**DOI:** 10.1101/2021.07.31.21261423

**Authors:** Steven J. Krieg, Carolina Avendano, Evan Grantham-Brown, Aaron Lilienfeld Asbun, Jennifer J. Schnur, Marie Lynn Miranda, Nitesh V. Chawla

## Abstract

COVID-19 remains a global threat in the face of emerging SARS-CoV-2 variants and gaps in vaccine administration and availability, and organizations must be prepared to detect and mitigate its risk to their people and activities. In this report we share key lessons learned from an adaptive COVID-19 testing program implemented at a mid-sized university in the Midwest. The program utilized two simple, diverse, and easily interpretable machine learning models to quickly and accurately predict which students were at elevated risk for contracting COVID-19 and should be called proactively for testing. Our adaptive testing cohorts produced positivity rates that were 26% higher than the random cohort: 0.58% positivity (95% CI 0.47% to 0.68%) from 19,171 tests, and 0.46% positivity (95% CI 0.41% to 0.51%) from 64,003 tests, respectively. Within 14 days of their selection, 2.94% of the adaptive cohort tested positive, compared to 1.27% of the random cohort. Close contacts who were predicted by the adaptive testing models received a COVID-19 test within an average of 0.94 days (95% CI 0.78 to 1.11) of the source testing positive, while those who were manually contact traced were tested in an average of 1.92 days (95% CI 1.81 to 2.02). These results suggest that machine learning strategies can improve surveillance testing effectiveness, especially in a university setting, by effectively distributing testing resources and potentially reducing community transmission.

## 1 Introduction

While schools, business, and other institutions seek to resume normal operations, the risk of SARS-CoV-2 and its emerging variants remains a global threat, especially as global vaccine rollouts remain in progress [6, 10]. These organizations must therefore be prepared to detect and mitigate its risk to their people and activities. In this report we share key lessons learned from an adaptive COVID-19 testing program implemented at a mid-sized university in the Midwest. The adaptive testing program utilized two different, data-driven network models to quickly and accurately predict which students had an elevated risk of contracting COVID-19 and should be called proactively for testing. Both models utilized a social network representation of the university community in which each node represented a person (our analysis focuses exclusively on students) and each edge represented a connection between two people (e.g., roommates, enrolled in the same course, active on the same sports team). The first model used this network to predict individual student risk via node risk propagation, and the second model predicted contact tracing relationships via multi-relational link prediction. The key difference between the two lies in the problem formulation: the first model was trained for a node-level task (classifying students as high-risk or low-risk), while the second was trained for an edge-level task (predicting contact tracing relationships between students). This difference resulted in models that were diverse and complementary, able to identify high-risk individuals and communities within the campus network while reducing the overhead of manual contact tracing. The success of this program suggests that machine learning strategies can improve the effectiveness of surveillance testing, especially in a university setting. Such approaches may help to more effectively distribute testing resources and reduce community transmission.

### 1.1 Adaptive Testing Overview

The adaptive testing program was one of many COVID-19 mitigation strategies implemented throughout the 2020-21 academic year at the university [1]. During the fall of 2020, 1,556 students (12.0%) and 200 faculty and staff tested positive from a total of 88,283 tests. In just the first four weeks of the spring 2021 semester (Feb. 3 through Mar. 2, 2021), another 734 students (5.7%) and 34 faculty and staff tested positive from a total of 57,661 tests. This provided a rich set of test results, contact tracing interviews, and symptom reporting to use as training data. The situation also necessitated urgent intervention—especially with respect to asymptomatic and presymptomatic cases, which contributed significantly to community transmission [5]. Thus a targeted and data-driven adaptive testing program was initiated on March 3, 2021 to *supplement* general surveillance testing, manual contact tracing, quarantine/isolation protocols, and self-reported health checks with more targeted and data-driven testing. While the program also included faculty and staff, our analysis focuses exclusively on students (undergraduate, graduate, and professional) as they constituted the majority of cases and testing appointments.

During each day of the program, two cohorts of students were sampled for surveillance testing. The first, which we call the general surveillance cohort, was determined by the students selecting a day of the week to be tested to ensure each student was tested once every week. The second, which we call the adaptive cohort, was selected via two distinct machine learning models—one trained on streaming data to predict SARS-CoV-2 transmission directly and one trained using data from the fall semester to predict contact tracing relationships. Using the set of active COVID-19 cases as input, both models generated a risk score for all other active students based on their connections to the active cases within the network representation of the university. Depending on the available testing resources (which increased substantially from fall to spring), the adaptive testing team chose the *n* students with the highest predicted risk from each model, with additional weight being given to students identified as high risk by both models, and added them to the adaptive cohort. Students who were already being tested for another reason such as reporting symptoms, living at the same address as a new positive case, having been contact traced, or having been tested twice already during the week (defined as a 7-day period from Monday to Sunday) were excluded from the adaptive sampling. Students in both the general surveillance and adaptive cohorts were notified of their selection for testing via email and text and administered either a saliva or nasal reverse transcription–polymerase chain reaction (PCR) test at the university testing center within 24 hours of notification.

## 2 Results

The adaptive testing program began on March 3 and concluded on April 30. Cohorts were tested daily with only a few exceptions (e.g., no adaptive tests were administered from April 17-19 to provide testing staff with time off during Easter weekend). During this period 97,604 total tests were administered to students: 64,003 (65.6%) to the general surveillance cohort, 19,171 (19.6%) to the adaptive cohort, and the remaining 14,430 (14.8%) to other cases such as students who reported symptoms or were contact traced. A total of 641 students tested positive: 297 (46.3%) during a general surveillance appointment, 111 (17.3%) during an adaptive testing appointment, and the remaining 235 (36.3%) during symptomatic appointments. The adaptive cohort thus produced a positivity rate of 0.58% (95% CI 0.47% to 0.68%), about 26% higher than the random cohort’s 0.46% positivity rate (95% CI 0.41% to 0.51%).

Many students returned for a follow-up test within several days of being selected for the adaptive cohort. When we look beyond the same-day test results, students selected for adaptive cohorts were even more likely to test positive. Within five days of being called for testing, 0.57% and 1.47% of the general surveillance and adaptive cohorts tested positive, respectively. As Figure 1 demonstrates, this gap in positivity rate between the two cohorts widens with the length of the follow-up window for at least 14 days after selection to the adaptive cohort.

**Figure 1:**
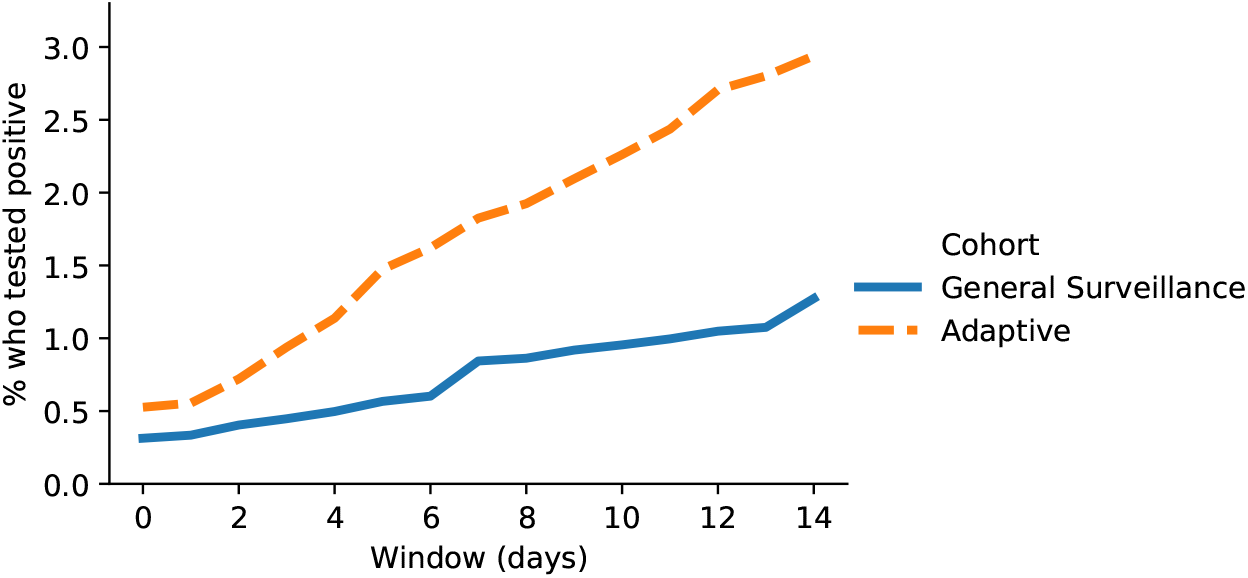
The percentage of students who tested positive from a follow-up test after being selected for one of the testing cohorts. Day 0 is the day they were selected for the cohort.

We also note that the adaptive cohort is essentially comprised of three groups of students: 10,251 who were selected by only the node risk (NR) model, 8,089 who were selected only by the link prediction (LP) model, and 2,608 who were selected by both models. As Figure 2 shows, students selected by both models were by far the most likely to test positive, producing a 0.8% positivity rate on the initial test, and with a total of 2.72% testing positive within five days. The students selected by both models tended to have the highest predicted risk from each model individually via connections like living on the same dorm floor or being in multiple courses with a student who had tested positive. Beyond these high-risk connections, we did not observe significant differences between the models with respect to the types of connections that produced high-risk predictions.

**Figure 2:**
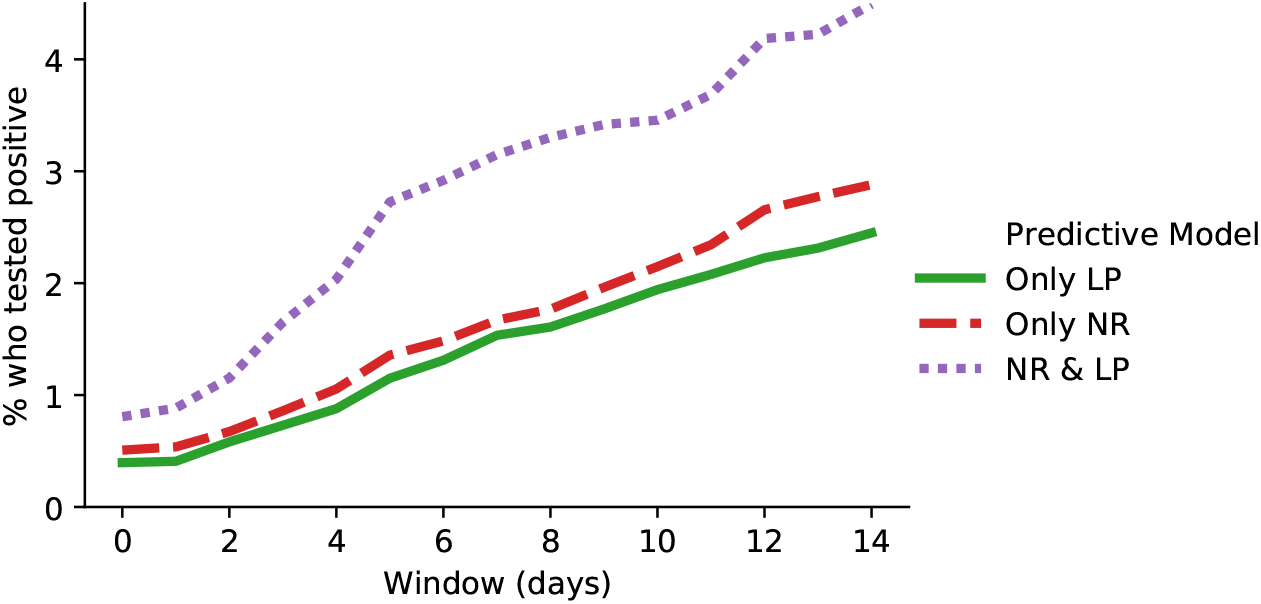
The percentage of students from the adaptive cohort who tested positive as predicted by the node risk (NR) and/or link prediction (LP) models. Day 0 is the day they were selected for the adaptive cohort.

Another key finding is that the adaptive testing program resulted in a significantly shorter average time to test for close contacts. Of 1,907 contacts that were traced to the 641 positive cases, 1,483 were administered a test on campus within seven days. 188 were administered a test via selection to the adaptive cohort within an average of 0.94 days (95% CI 0.78 to 1.11), while the remaining 1,295—who were tested only after being notified of their exposure by contact tracers or the student who exposed them—were administered a test within an average of 1.92 days (95% CI 1.81 to 2.02). Figure 3 shows the full distribution of test timings for confirmed contacts.

**Figure 3:**
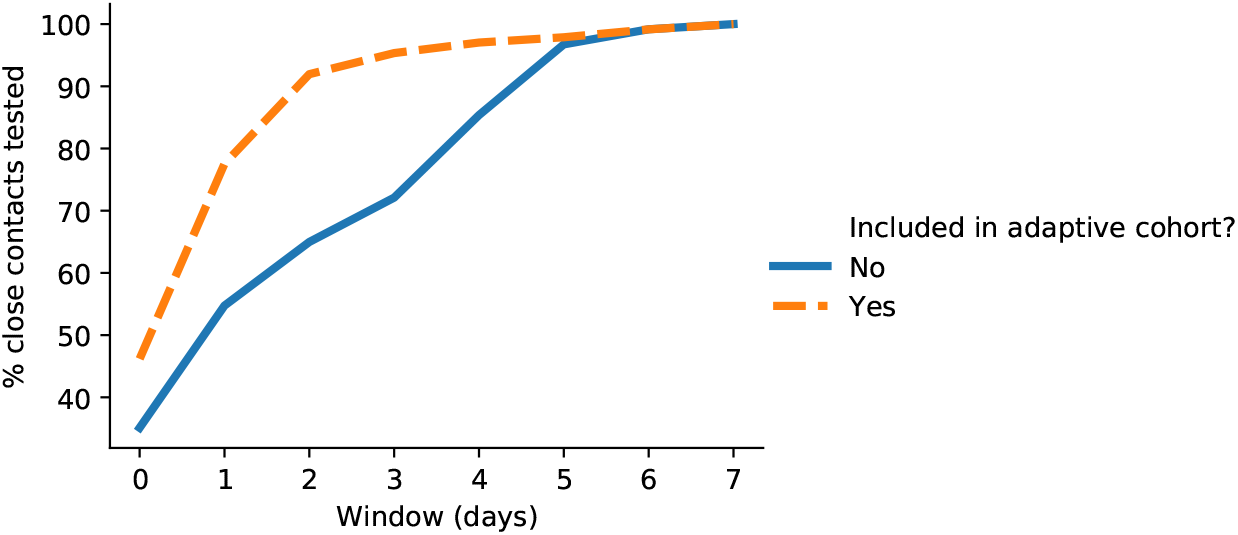
The distribution of average time to receive a test for students who were exposed to COVID-19 via close contact with another student. Day 0 is the day the exposing student tested positive.

We additionally found that the average same-day response rates were 78.1% and 95.0% for the general surveillance and adaptive cohorts, respectively. While the general surveillance cohort was least likely to respond on Saturdays and Sundays (61.2%), only 8.3% of these appointments were scheduled on these days, and the response rate for weekday appointments was only 79.6%. The daily distribution of appointments for the adaptive cohort, on the other hand, was relatively uniform: 23.8% were scheduled on a Saturday or Sunday with a 95.8% response rate. While undergraduate students produced higher response rates than graduate and professional students, the differences were marginal (79.4%, 78.1%, and 82.1%, respectively) for the general surveillance cohort. For the adaptive cohort the differences between means were more significant (96.8%, 85.6%, and 92.7%, respectively); however, the number of adaptive tests administered to graduate and professional students was only 442 (2.3%) and 631 (3.3%), respectively. In any case, the response rates for adaptive testing were consistently higher than for general surveillance.

## 3 Discussion

Rapidly identifying COVID-19 cases is of paramount importance to reducing community transmission [7]. While the adaptive program did produce higher same-day positivity rates, the model predictions gained value over time (Figure 1). We suggest that this is explained largely by the following two factors. First, the incubation period of SARS-CoV-2 means that students who are called for adaptive testing immediately (the next day) after a close contact tests positive may not have built enough of a viral load to be detected by the PCR test [11]. Second, networks excel at modeling transmission dynamics in local communities, i.e. transmission in a dorm floor. High-risk individuals should therefore be tested as soon as possible, but a single negative test does not allay the risk of further transmission through a third party. An ideal follow-up protocol should include cadenced re-testing for at least 14 days.

Prior to March 3, students were informed of the adaptive testing program and its data-driven approach to identifying individuals who were at high-risk. Therefore, some of the differences in response rates could be due to students’ perceiving an adaptive testing call as more important than a general surveillance one. However, it is also possible that adaptive testing appointments were correlated with individuals’ knowledge that they had been potentially exposed. It seems likely that both of these factors contributed to the increased response rate.

The shortcomings of contact tracing have been well-documented, including that the process requires a high amount of manual effort [4] and that individuals can be reluctant to disclose their social activity [2]. The ability of an adaptive testing program to identify high-risk individuals and produce shorter times to test for close contacts can mitigate both of these problems. However, while our models are an effective supplement to manual contact tracing, they are not a replacement for it. There are many close contacts that were not predicted by our models, and many student behaviors that are not captured in a social network.

The simplicity of the adaptive testing models lends itself well to implementation by others. Universities can easily construct a social network representation using similar data. Other organizations could likewise create a social network from reporting and departmental relationships, meeting attendees, or office layouts. We experimented with other approaches and data, most notably using the network to model the spread of symptoms rather than just positive cases; however, we found that student symptom profiles were too inconsistent to effectively predict viral transmission [5]. The fact that the link prediction model performed similarly to the node risk model (Figure 2) suggests that predicting contact tracing relationships is an effective proxy for predicting viral transmission. This observation can be utilized to encourage diversity by training models for different tasks. Specifically, COVID-19 testing produces node-level data and therefore support node-level tasks; likewise, contact tracing produces edge-level data and thus supports edge-level tasks. As our results demonstrate, training models for both tasks can make predictions that are diverse and complementary. However, even in the absence of data on positive tests, organizations could still collect and use data on known close contacts to train a predictive model. Such approaches may prove to be necessary to rapidly detect positive cases and drive down community transmission.

## 4 Materials and Methods

Foundational to the adaptive testing program was the modeling of the university as a heterogeneous network, a widely-used formalism in graph theory and network analysis [9]. Formally, we define a network *G* = (*V, E*), where *V* is a set of *n* nodes and *E* is a set of *m* edges. Each *u* ∈ *V* represents one student, and each *e* ∈ *E* is a tuple (*u, v, t*) that represents a relation between two nodes *u* and *v* of type *t*. Possible relation types included two nodes sharing the same home or dorm address, being enrolled in the same course, playing the same team sport, sharing a dorm floor or building, and being confirmed as close contacts by a contact tracer. We additionally consider a weight function *w* : *E* → ℝ that maps each edge to a real-valued weight, or 0 if the edge does not exist. In our context, all edges have a weight of 1 except for students who were enrolled in the same course(s), in which case the edge weight was the number of courses they shared. This representation is thus a flexible and expressive means of modeling interactions among students in the context of a large community, and served as the input to the predictive models.

### 4.1 Predictive Modeling

In designing the the predictive models we prioritized the following principles:

1. Simplicity. The operational needs of the program were urgent, so we designed models that were relatively simple to develop, test, and deploy. This is true of both the learning algorithms and the underlying data.
2. Diversity. The combination of diverse models is the cornerstone of the success of ensemble methods in machine learning [14]. In our case, we encouraged diversity by optimizing each model for a different task within the campus social network: one model for a node-level task, and the other for an edge-level task. Taken together, the strengths and limitations of both models proved to be complementary in solving the operational problem.
3. Interpretability. The outputs of the predictive models were monitored by the adaptive testing team and subject to further operational constraints. For example, roommates of students who tested positive were already being called for testing, and so were excluded from adaptive testing selection. However, roommate connections contribute significantly to the campus social network structure. By designing models with interpretable outputs, we simultaneously enabled the models to make full use of the information provided by roommate connections and the adaptive testing team to make informed decisions with the aid of model outputs.

#### 4.1.1 Node Risk Prediction

The first model predicted risk at the node level by first assigning each node *u* ∈ *V* a base risk score *P*_0_ based on whether the student had recently tested positive, been assigned to quarantine/isolation, or reported COVID-19 symptoms. Next, each node sent a portion of its risk to its neighbors. This risk propagation approach, known more generally as message passing, is foundational to many network inference tasks [8]. For a given node *u* we iteratively computed its final risk *P*_*i*_(*u*) according to the following:

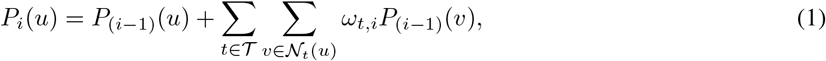

where *i* is a parameter defining the number of message passing iterations, *𝒯* is the set of edge types, 𝒩_*t*_(*u*) is the set of *u*’s neighbors via edge type *t* (i.e., (*u, v, t*) ∈ *E* for all *v* ∈ 𝒩_*t*_(*u*)), and *ω*_*t,i*_ is a learned weight parameter associated with edge type *t* at hop *i*. Intuitively, this means that during each iteration each node’s risk is updated with the weighted sum of the risk of its neighbors, where the weights are fixed for each combination of edge type and hop number. We set *i* = 2 such that each node’s final risk is influenced by nodes up to two hops away in the network.

To learn the set of all weights *ω*_*t,i*_, we utilized the following evolutionary strategy:

1. Given an initialized set of weights, target a random previous day in the semester, denoted as *d*.
2. Create a copy of the weights and make several small and random adjustments to them.
3. Simulate the testing results for day *d* via Equation 1, and evaluate the predictions for both sets of weights.
4. Keep the weight set that more accurately predicted which nodes tested positive on day *d*.
5. Repeat steps 1-4 until convergence.

This approach was simple and interpretable (as compared to state-of-the-art models like graph neural networks [12]), and allowed us to easily include new testing results as they became available during the semester (however, in order to minimize bias we utilized general surveillance and symptomatic test results, not adaptive testing results). To choose students for the adaptive cohort, we simply selected the nodes with the highest values of *P*_*i*_ that were susceptible (i.e., had not yet tested positive).

#### 4.1.2 Link Prediction

The second model predicted risk at the edge (link) level by utilizing correlations between edge types to predict unobserved contact tracing relationships. Our approach to this problem, known as multi-relational link prediction, is adapted from the work of Yang *et al*. [13]. For each pair of nodes *u, v* ∈ *V* we compute the probability of a contact tracing relationship *P* (*u, v*) according to:

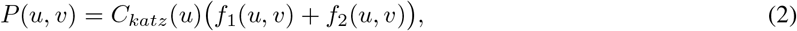

where *C*_*katz*_(*u*) is the Katz centrality of node *u* [3], and *f*_1_ and *f*_2_ represent the one-hop and two-hop information flow, respectively, from *u* to *v*. We define *f*_1_ as follows:

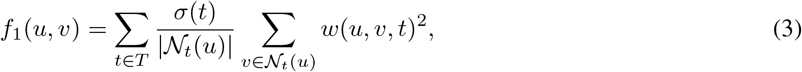

where *σ*(*t*) is a learned conditional probability that two nodes *u* and *v* will be contact traced given that they are connected by an edge of type *t*. We compute *f*_2_ in the same manner as *f*_1_, but with respect to a two-hop neighbor graph of *G*. The 2-hop neighbor graph is constructed by adding an edge of type *t* between any two nodes *u* and *v* if they are both neighbors to a common node *x* via edge type *t*. To compute the conditional probabilities for *σ*, we utilized a training network built from student and contact tracing data from the (previous) fall 2020 semester. This approach assumes that although student information (dorm address, course schedule, etc.) may change between semesters, the conditional probability distribution of contact tracing relationships do not.

To choose students for the adaptive cohort, we first computed *P* (*u, v*) for each node *u* that had tested positive within the previous four days with respect to each other node *v ≠ u*. Then we chose the nodes that were most likely to be contact traced to a positive node *u* that were also susceptible (i.e., had not yet tested positive).

## Data Availability

The data and code for this study are available from the corresponding author upon reasonable request.

